# Effects of weather extremes on fecal contamination along pathogen transmission pathways in rural Bangladeshi households

**DOI:** 10.1101/2023.12.27.23300582

**Authors:** Caitlin Niven, Mahfuza Islam, Anna Nguyen, Andrew Mertens, Amy J. Pickering, Laura H. Kwong, Mahfuja Alam, Debashis Sen, Sharmin Islam, Mahbubur Rahman, Leanne Unicomb, Alan E. Hubbard, Stephen P. Luby, John M. Colford, Benjamin F. Arnold, Jade Benjamin-Chung, Ayse Ercumen

## Abstract

**Background:** Weather extremes are predicted to influence pathogen exposure but their effects on specific fecal-oral transmission pathways are not well investigated. We evaluated effects of extreme rain and temperature during different antecedent periods (0-14 days) on *E. coli* along eight fecal-oral transmission pathways in rural Bangladeshi households.

**Methods:** *E. coli* was enumerated in mother and child hand rinses, food, stored drinking water, tubewells, flies, ponds, and courtyard soil using IDEXX Quanti-Tray/2000 in nine rounds over 3·5 years (n=26,659 samples) and spatiotemporally matched to daily weather data. We used generalized linear models with robust standard errors to estimate *E. coli* count ratios (ECRs) associated with extreme rain and temperature, defined as >90^th^ percentile of daily values during the study period.

**Findings:** Controlling for temperature, extreme rain on the sampling day was associated with increased *E. coli* in food (ECR=3·13 (1·63, 5·99), p=0·001), stored drinking water (ECR=1·98 (1·36, 2·88), p<0·0005) and ponds (ECR=3·46 (2·34, 5·11), p<0·0005), and reduced *E. coli* in soil (ECR=0·36 (0·24, 0·53), p<0·0005). Extreme rain the day before sampling was associated with reduced *E. coli* in tubewells (ECR=0·10 (0·02, 0·62), p=0·014). Effects were similar for rainfall 1-7 days before sampling and slightly attenuated for rainfall 14 days before sampling.

Controlling for rainfall, extreme temperature on the sampling day was associated increased *E. coli* in stored drinking water (ECR=1·49 (1·05, 2·12), p=0·025) and food (ECR=3·01 (1·51, 6·01), p=0·002). Rainfall/temperature was not consistently associated with *E. coli* on hands and flies.

**Interpretation:** In rural Bangladesh, measures to control enteric infections following weather extremes should focus on reducing contamination of drinking water and food stored at home and reducing exposure to surface waters.

**Funding:** Bill & Melinda Gates Foundation, National Institutes of Health, World Bank.

**Research in Context:** *Evidence before this study:* Higher temperatures and levels of rainfall are associated with increased waterborne and vector-borne disease incidence, including child diarrhea. However, the specific pathways that facilitate increased transmission of diarrheagenic pathogens under these weather conditions have not been well investigated. We searched Google Scholar for articles published since 2000 using the following search terms: (“climate change” OR weather OR temperature OR heatwave OR rainfall OR precipitation) AND (pathogen OR enteropathogen OR “*Escherichia coli*” OR *“E. coli”* OR “fecal indicator” OR “fecal contamination”) AND (environment OR water OR hands OR food OR soil OR flies). A large body of literature has evaluated the effect of rainfall or temperature on water quality and generally found that higher temperatures and magnitudes of rainfall were associated with higher levels of fecal indicator bacteria, such as *Escherichia coli* (*E. coli*), in surface and groundwaters, public and private drinking water sources and drinking water stored at homes. However, studies on the impact of rainfall and temperature on fecal contamination along non-waterborne fecal-oral transmission pathways are limited. Contamination of food stored at home has been linked to storage temperature. We found only one study on hand contamination with respect to weather, which found lower *E. coli* counts on child hands when daily temperatures were higher but no effect from rain. No studies have simultaneously assessed the effects of weather events on a comprehensive set of fecal-oral transmission pathways, which are typically described with the F-diagram and can include drinking water (at the source or stored), surface waters, caregiver and child hands, food, soil and flies.

*Added value of this study:* We spatiotemporally matched historical meteorological data to over 26,000 *E. coli* measurements collected in nine rounds over 3·5 years in a randomized controlled trial in rural Bangladesh. *E. coli* was measured across eight different pathogen transmission pathways in the domestic environment. To our knowledge, this study is the first to utilize a large longitudinal dataset of environmental measurements collected over multiple years to investigate how increased rainfall and temperature affect fecal contamination across the full span of pathways described by the F-diagram. Our findings can help identify fecal-oral transmission pathways that are the most susceptible to extreme weather events and should be prioritized for intervention in their wake, as well as offer guidance on the time windows when interventions should be implemented with respect to weather events to interrupt these pathways in the context of climate change. This study can inform the effective delivery of WASH interventions, supporting climate change adaptation to reduce the enteric disease burden associated with weather extremes.

*Implications of all the available evidence:* Extreme rainfall within two weeks of sampling was associated with increased *E. coli* contamination in stored food, stored drinking water and ponds, and reduced contamination of tubewell water and courtyard soil. Extreme temperature during the same timeframe was associated with increased contamination of stored food and stored drinking water. These findings illuminate environmental mechanisms behind previously reported increases in diarrheal diseases associated with extreme rainfall and temperature. Our findings suggest that, as extreme weather events become more common with climate change, intervention efforts to control exposure to fecal contamination in rural Bangladesh should prioritize reducing contamination of stored food and drinking water as well as reducing exposure to contaminated surface waters.

## Introduction

Studies in low– and middle-income countries (LMICs) indicate strong associations between increased temperature/rainfall and child diarrhea but the specific mechanisms behind these associations are not well investigated^1–3^. Diarrhea-causing pathogens are fecal-orally transmitted through environmental compartments (e.g., water, soil), food, fomites, and vectors (e.g., flies), which can all be influenced by weather^4,5^. Increased rainfall leads to greater mobilization and loading of pathogens into waterbodies, soil and crops while higher temperatures can support viability, reproduction and incubation in the environment, both for vectors and pathogens^2,6–8^.

Increased fecal indicator levels in drinking water sources are observed following higher rainfall and temperature in LMICs^9^. Additionally, proliferation of fecal microorganisms in food is dependent on storage temperature^10^. In a study in Bangladesh, *E. coli* levels in food stored at home increased with each 1°C increase in the storage area temperature^11^. One study in Korea found increased prevalence of pathogenic *E. coli* on raw vegetables during months with higher temperatures while a laboratory study did not find temperature effects on bacterial load on vegetables^12,13^. Data on weather effects on other fecal-oral transmission pathways are scarce. A single study to date has found lower *E. coli* counts on children’s hands following increased temperature but not heavy rainfall in Kenya^14^.

Bangladesh is one of the world’s top ten most climate-vulnerable countries due to its microcosm of climates, proximity to the Himalayas, and unique hydrogeology as a delta and floodplain^15^.

The region experiences multiple climate-related issues such as saltwater intrusion, flooding from glacial melting, water and food insecurity, and shifts in infectious disease patterns^15^. Studies in Bangladesh have shown increased occurrence of infectious diseases during the monsoon season, and higher prevalence of pneumonia, diarrhea, and infections with enteropathogens associated with increased rainfall^3,7^. Heavy rainfall and high temperatures have also been associated with increased fecal contamination of groundwater from tubewells in Bangladesh, while the monsoon season has been associated with higher levels of *E. coli* in soil, ponds, tubewell water, stored drinking water, food, child hands and flies^16,17^. However, the latter study defined the monsoon season based on calendar month rather than location-specific rainfall data. Climate change may influence both the timing and duration of monsoon seasons, further supporting the use of data-driven definitions of weather events^18^.

Here, we aim to assess the effects of extreme rainfall and temperature on *E. coli* contamination in the domestic environment, utilizing environmental samples and spatiotemporally matched daily weather data from rural Bangladesh.

## Methods

### Study design

We used data from the WASH Benefits randomized controlled trial in rural Bangladesh (NCT01590095), which measured the effects of water, sanitation, hygiene (WSH) and nutrition interventions on child diarrhea and growth^19^. The trial enrolled 5,551 pregnant women and followed their birth cohort (index children) for two years. Six to eight enrolled households formed a study cluster, and eight adjacent clusters formed a block. Within each block, clusters were randomized to intervention vs. control arms. Details of the study design have been previously published^20^.

### Environmental data

Environmental samples were collected from the control, sanitation intervention and combined WSH intervention arms longitudinally in nine rounds over 3·5 years. Analyses in this study pooled data across these study arms. Sampled pathways included child and mother hand rinses, stored food for children <5 years (primarily rice), stored drinking water, source water (groundwater from tubewells), ponds, courtyard soil and captured flies. Child hand rinses and stored drinking water were collected during all rounds, mother hand rinses during eight rounds, soil and food during three rounds, and tubewell, pond and fly samples once.

Samples were processed at the field laboratory of the International Centre for Diarrhoeal Disease Research, Bangladesh (icddr,b) with IDEXX Quanti-Tray/2000 using Colilert-18 media and incubated at 44·5°C^21^. We enumerated the most probable number (MPN) of *E. coli* per 100 mL of source/stored water and pond samples, per two hands for hand rinses, per dry gram for food and soil, and per fly. Values for non-detects were imputed as half the lower detection limit (0·5 MPN) and values above the upper detection limit (2419·6 MPN) were imputed as 2420 MPN; detection limits per reporting unit differed by sample type due to different dilution factors and, for food and soil samples, varying moisture content (Table S1). Sample collection and processing details have been previously reported^22,23^.

### Weather data

Daily rainfall and temperature observations were obtained from GloH2O’s Multi-Source Weighted-Ensemble Precipitation (MSWEP) dataset with a 3-hour 1·0° resolution and the National Aeronautics and Space Administration’s Famine and Early Warning Systems Network (FEWS NET) Land Data Assimilation System for Central Asia (FLDAS-Central Asia) dataset with a daily 0·01° resolution using our study area’s bands of latitude (23·7-25·0) and longitude (89·8-90·9)^24–26^. Missing data were imputed using available data from the nearest neighboring location. Daily weather data were matched to *E. coli* data based on the sample collection date and the GPS coordinates of study households using nearest neighbor matching, and a Cartesian distance was calculated for each selected match.

We generated weather variables for antecedent periods of 0, 1, 2, 7 and 14 days before sample collection. For each period, we calculated the rolling average rainfall (mm) and temperature (°C). We used daily maximum rainfall and temperature values to tabulate whether heavy/extreme rainfall, extreme heat, or a heatwave occurred during that window. Heavy and extreme rainfall were defined as ≥80^th^ and ≥90^th^ percentile, respectively, of daily rainfall values for our study region over the study period^2,6,7^. Extreme heat was defined as ≥90^th^ percentile of daily temperature values, and a heatwave as daily maximum temperature >95^th^ percentile for three consecutive days^27^. For a sensitivity analysis, we defined elevated heat as ≥80^th^ percentile of daily temperature values. We created categorical variables for rainfall intensity (no, some, heavy, extreme) and temperature (below median, above median, extreme/elevated) for each antecedent period and binary variables for heatwave occurrence for the 7-day and 14-day periods.

### Statistical analysis

We used generalized linear models (GLM) with a negative binomial error distribution and robust standard errors to estimate *E. coli* count ratios (ECRs) for periods with different rainfall intensities vs. no rainfall, periods with extreme/elevated and above-median temperature vs. below-median temperature, and periods with vs. without a heatwave, separately by sample type. Models for rainfall controlled for rolling average temperature during the same period, and vice versa. Models also controlled for intervention status (binary variable for sanitation/WSH intervention vs. controls) and covariates expected to influence *E. coli* levels, measured either at the trial’s baseline (mother’s age and education, number of children <18 years in the household, number of individuals living in the compound, food security, asset-based wealth index, wall and floor materials, drinking water source, minutes to primary water source, and number of cows, goats, and chickens) or at the time of sample collection (sex and age of index child). Three additional variables were included for stored drinking water models (duration of storage and whether the storage container was covered and had a narrow mouth), and one additional variable was included for food models (hours since the food was prepared), due to their relevance to these sample types. For stored water and food samples, we also tabulated *E. coli* levels under different storage and weather conditions to identify any protective effects from safe storage practices. Analyses were conducted in R (version 4.1.2, RStudio 2022.02.1+461).

## Results

Field staff visited 1840 households in the control, sanitation and WSH arms once between July 2013-March 2014, and 720 households in the control and sanitation arms eight times between June 2014-December 2016 to collect samples, resulting in 652 study dates and 7253 study observations (unique combinations of household GPS coordinates and date). We collected 6350 stored water, 2181 stored food, 5397 mother hand rinse, 7092 child hand rinse, 1669 source water, 2538 soil, 822 pond, and 610 fly samples (Table S2). Geometric mean *E. coli* counts were 7·8 MPN/100 mL in stored drinking water, 5·0 MPN/g in food, 29·5 MPN/two hands on mother hands, 20·5 MPN/two hands on child hands, 0·9 MPN/100 mL in source water, 125,047 MPN/g in soil, 5397 MPN/100 mL in ponds, and 712 MPN per fly (Table S4).

Rainfall data were available for all study observations. The mean distance between study households and the location of rain measurement was 0·4 km (range: 8.2 m-0.76 km). The temperature dataset had 0·3% (2/652) missing daily values, which were imputed using the nearest available temperature data for these two dates. Daily rainfall ranged between 0–157·1 mm over the study period. The 80^th^ percentile (heavy rainfall) corresponded to 16·4 mm, and the 90^th^ percentile (extreme rainfall) to 28·2 mm. Daily temperature ranged between 13·1–34·2°C. The 90^th^ percentile (extreme temperature) corresponded to 30·2°C and the 95^th^ percentile (heatwave) to 30·9°C. Within 14 days before sampling, 32·7% (2373) of study households experienced extreme rainfall and 27·3% (1984) experienced extreme temperature. A heatwave occurred within the past 14 days for 12·5% (910) of study observations (Table S3). Heaviest rain and extreme temperatures occurred between the months of March and August (Figure S1).

Controlling for temperature and potential confounders, extreme rainfall on the day of sampling was associated with 2 to 4-fold higher *E. coli* counts in stored water (ECR=1·98 (1·36, 2·88), p<0·0005), food (ECR=3·13 (1·63, 5·99), p=0·001) and ponds (ECR=3·46 (2·34, 5·11), p<0·0005), and lower *E. coli* counts in soil (ECR=0·36 (0·24, 0·53), p<0·0005) compared to no rainfall (Figure 1, Table S5). Extreme rainfall the day before sampling was associated with lower *E. coli* counts in tubewells (ECR=0·10 (0·02, 0·62), p=0·014) (Figure 1, Table S5). For most pathways, effects were similar when rainfall occurred 1-7 days before sampling and attenuated when it occurred 14 days before sampling (Figure 1, Table S5). Increasing rainfall appeared to be associated with higher *E. coli* counts in/on flies but effects were not consistent across different rainfall intensities and antecedent periods (Figure 1, Table S5). There was no consistent association between rainfall and *E. coli* on child or mother hands (Figure 1, Table S5).

**Figure 1.**
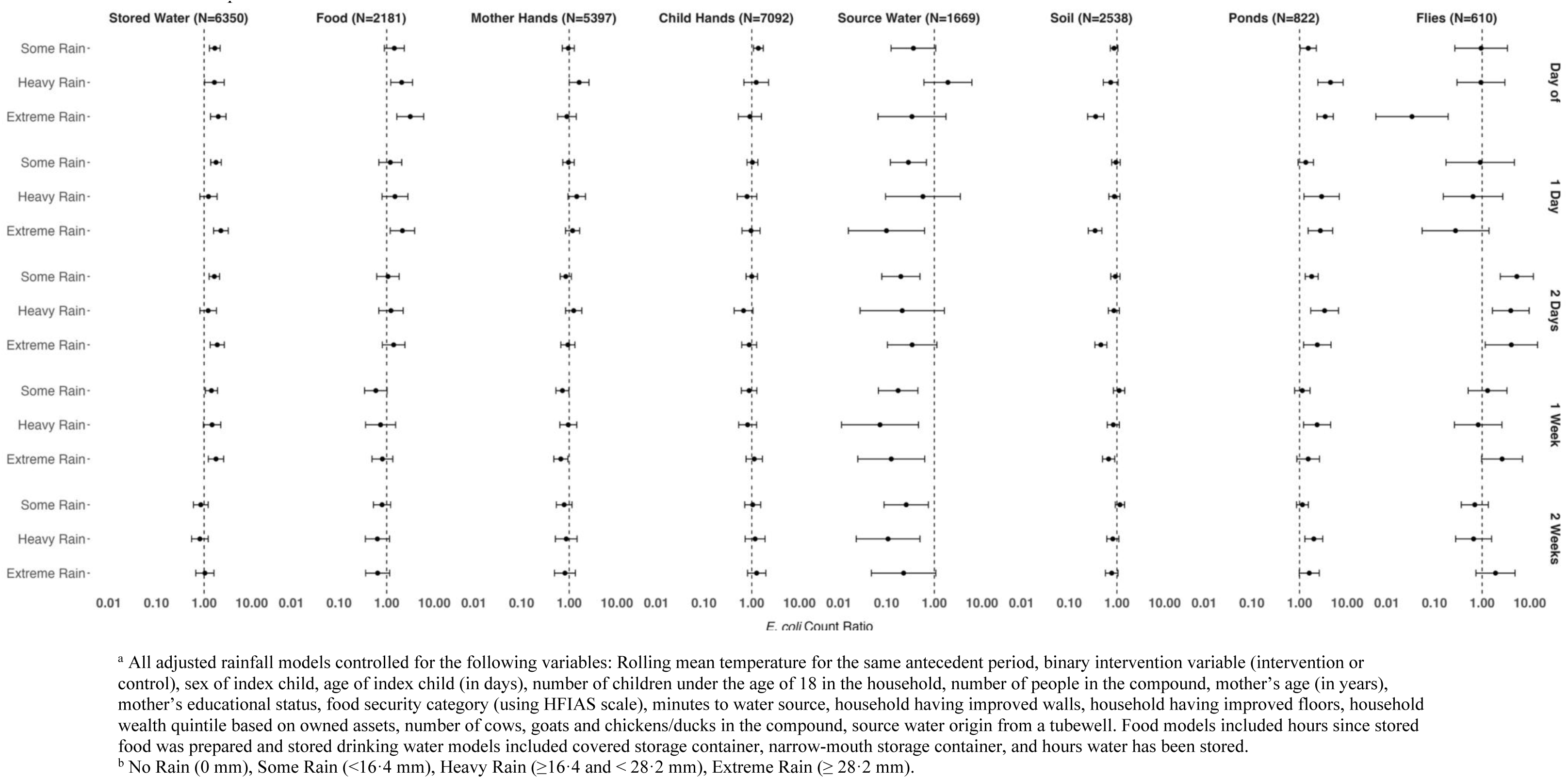
Adjusted^a^ *E. coli* count ratios by sample type associated with rainfall intensity^b^, compared to no rainfall, during different antecedent periods.

Controlling for rainfall and potential confounders, extreme temperature on the day of sampling was associated with higher *E. coli* counts in stored drinking water (ECR=1·49 (1·05, 2·12), p=0·025) and food (ECR=3·01(1·51, 6·01), p=0·002). Effects on stored water and food were similar for all antecedent periods (0-14 days) (Figure 2, Table S5). Extreme temperature had no effect on child or mother hands across any antecedent period but above-median temperature 0-2 days before sampling was borderline associated with increased *E. coli* on child hands (Figure 2, Table S5). We could not estimate effects of extreme temperature on tubewell water, ponds, and flies because only a small number of sampling days for these sample types fell on periods with extreme temperature. The sensitivity analysis using elevated temperature showed no effects on tubewell water, ponds, and flies, and effects similar to extreme temperature on stored water and food (Figure S2). A heatwave within 7 or 14 days before sampling was associated with lower *E. coli* counts in soil (ECR=0·54 (0·38, 0·78), p=0·001 and ECR=0·50 (0·36, 0·70), p<0·0005, respectively) but had no significant effect on other pathways (Figure S5). The impacts of a heatwave could not be estimated for tubewell water, ponds, and flies due to data sparsity.

**Figure 2.**
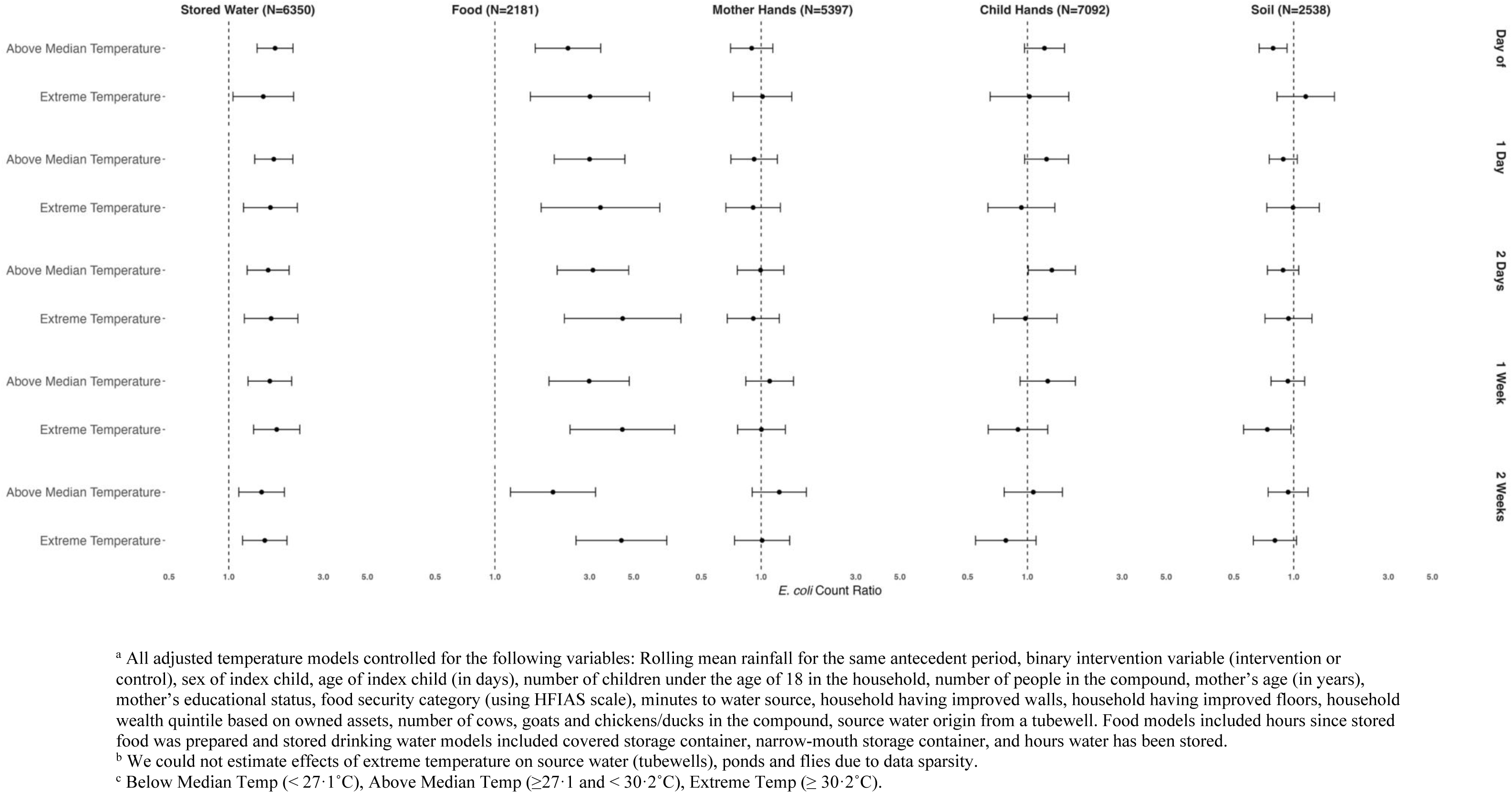
Adjusted^a^ *E. coli* count ratios by sample type^b^ associated with above-median and extreme temperature^c^, compared to below-median temperature, during different antecedent periods.

Among drinking water storage containers, 45% had a narrow mouth and 30% were covered. At the time of sample collection, water had been stored for a median of two hours (interquartile range (IQR)=1-4, range=0-600). Mean *E. coli* counts were 123 MPN when water was stored for <1 hour, 148 MPN when stored for 1-8 hours and 222 MPN when stored for ≥8 hours. Water stored for ≥8 hours had a mean *E. coli* level of 133 MPN if stored in a covered container and 475 MPN if stored in an uncovered container on rainy days, compared to 69 MPN if stored in a covered container and 206 MPN if stored in an uncovered container on a day with no rain.

The median food storage time was 3·5 hours (IQR=2-5, range=0-29). Mean *E. coli* counts were 157 MPN for food stored <4 hours, 346 MPN for food stored between 4–8 hours, and 2467 MPN for food stored for ≥8 hours. The ambient temperature on food sampling days was between 16·7– 33·3°C, within the “danger zone” for ideal bacterial incubation in food (5–60°C) and close to the optimum growth temperature (around 35°C) for enteric bacteria^28^. We found that 74% of food samples were stored for ≥2 hours (maximum recommended storage duration at room temperature (29)) on days where the ambient temperature was equal to or exceeded 20°C; these samples had a mean *E. coli* level of 645 MPN vs. 46 MPN for samples stored for <2 hours in the same conditions. Also, 0·3% of food samples were stored for ≥1 hour (maximum recommended storage duration at ambient temperatures >32°C on days where the temperature was at or above 32°C^28^; these samples had a mean *E. coli* level of 1297 MPN while no food samples were stored for <1 hour in these conditions to allow comparison.

## Discussion

Extreme rainfall and temperature occurring up to two weeks before collection of environmental samples significantly affected *E. coli* contamination along various fecal-oral transmission pathways in rural Bangladeshi households. Extreme rainfall was associated with increased contamination of stored food, stored drinking water and ponds, and reduced contamination of tubewell water and courtyard soil. Extreme temperature was associated with increased contamination of stored food and stored drinking water. Associations were more pronounced when these weather extremes occurred within 7 days of sampling and slightly attenuated when they occurred 14 days before sampling. Heatwaves, both within 7 and 14 days prior to sampling, were associated with decreased soil contamination.

Samples collected from the ambient outdoor environment – surface waters (ponds), groundwater (tubewells), soil – were sensitive to rainfall but generally not temperature in our study and were influenced in different directions by rainfall. Increased contamination in ponds after rainfall is consistent with a previous study in Kenya that found higher levels of *E. coli* in river waters following rainfall, indicating flushing of environmental pathogens into waterbodies^29^. This is supported by the lower *E. coli* levels in soil we observed following extreme rainfall, suggesting that contaminants were diluted and/or flushed out of the soil environment with rain^6^. Rainfall was associated with reduced *E. coli* in tubewells, potentially indicating dilution from surface recharge. It has been hypothesized that contaminants flush into groundwater aquifers at the beginning of wet seasons, become diluted over the wet season with increased volumes of groundwater, and become concentrated in the dry season, while individual rainfall events cause rapid spikes in contamination followed by a decline^30^. A recent literature review found generally higher levels of *E. coli* in groundwater with increasing rainfall^9^. Our findings of reduced groundwater contamination 1-14 days following rainfall, despite increased contamination of surface waters during the same window, indicates that the surface recharge was effectively filtered in subsurface. Soils in our study region are primarily alluvial and silt; these soil types typically pose an effective barrier against pathogen transport^16,31^. Groundwater in regions with different hydrogeological features may experience different effects from rainfall. Also, tubewells are an improved water source; groundwater from unimproved or unprotected sources may be more vulnerable to contamination from runoff during heavy rainfall. Among samples collected from the outdoor environment in our study, the only consistently observed temperature effect was reduced *E. coli* in soil following heatwaves, consistent with prior evidence of bacterial die-off after prolonged heat exposure^32^.

For samples stored in the indoor home environment – stored drinking water and prepared food – both extreme rainfall and temperature were associated with increased contamination. Given that weather extremes did not adversely affect groundwater quality at the source, increased contamination of stored water likely stemmed from bacterial intrusion into storage containers and bacterial growth during collection, transport and/or storage following extreme rainfall and temperature^17,33^. *E. coli* levels increased with storage duration while samples from covered containers had less contamination than those from uncovered containers, both on days with and without rainfall. A previous study in Bangladesh found that safe storage of tubewell water in a container with a narrow mouth, tight-fitting lid and spigot significantly improved stored water quality and reduced diarrhea^34^. In a recent study in Kenya, heavy rainfall and extreme temperature were associated with increased *E. coli* in source water from boreholes, springs, and wells while heavy rainfall but not extreme temperature was associated with increased *E. coli* in stored drinking water^14^. In the same study, heavy rainfall did not increase stored water contamination among households that treated their water^14^. Our findings suggest that treatment and safe storage of drinking water is especially important following extreme rainfall and temperature. Household-level water treatment typically has low uptake; focused recommendations for treatment during these high-risk times may help achieve health benefits.

Similarly, higher *E. coli* counts in food may be due to increased bacterial growth during storage under rainy/hot conditions, consistent with prior evidence linking higher food storage temperature with increased contamination^10,11^. In our study, food was typically stored in a covered container and inside a cabinet or elevated from the ground^22^. Previous studies indicate substantial increases in fecal contamination in foods stored for >4 hours after preparation^11,35^.

The WHO recommends that food should not be stored for more than two hours at room temperature (20–22°C in settings with heating and cooling, unlikely in our study setting), and not be left out for more than one hour when ambient temperatures exceed 32°C^28^. In our study, *E. coli* levels doubled in food stored for an hour or more on days when the ambient temperature was ≥32°C compared to food stored for the same amount of time while the ambient temperature was 20°C. Our findings emphasize the importance of minimizing the duration of food storage during extreme ambient temperatures to reduce pathogen growth.

We did not find a consistent association between rainfall/temperature and contamination of child or mother hands. In the aforementioned recent study in Kenya, *E. coli* counts were lower on child hands on days with higher temperatures while there was no association with rainfall^15^.

Others have hypothesized that children may have cleaner hands during higher rainfall and temperatures because they may spend less time playing outside and/or have more water available for handwashing^14,36^. Children in our study were young (85% ≤3 years, 30% ≤1 year). Children in this age range have frequent hand contact with objects, indoor surfaces and floors; unmeasured contamination from these pathways may have obscured effects of ambient conditions. We are unaware of previous studies on associations between weather and caregiver hand contamination. *E. coli* levels on caregiver hands are highly temporally variable^36,37^; our findings indicate that the contribution of weather to contamination of mother hands may be negligible compared to other dominant factors (e.g., domestic chores, contact with domestic animals). Additionally, while others have found that caregiver hand contamination is a predictor of stored water contamination, our findings indicate that the higher *E. coli* levels we observed in stored drinking water and food after increased rainfall/temperature are not due increased mother or child hand contamination^37^.

This study spatiotemporally matched daily weather data to >26,000 *E. coli* measurements across eight fecal transmission pathways. Study strengths include a large sample size, longitudinal measurements over multiple years capturing a range of weather conditions, comprehensive set of fecal-oral pathways and use of daily weather data as opposed to regional definitions of seasonality. Our study also had limitations. The nearest available rainfall data were located up to 0·76 km from households and temperature data were aggregated over pixels covering a 0·01° by 0·01° area, potentially not reflecting the exact conditions at sampling sites. Any misclassification of weather conditions would be non-differential with respect to our outcome measures and therefore bias our findings towards the null. We measured fecal indicator bacteria; therefore, the behavior of pathogens, especially nonbacterial pathogens, may not be represented. For example, higher temperatures are associated with reduced viral infections and increased protozoan infections^1,38–40^. Future research should investigate weather effects on specific pathogens in the environment. However, *E. coli* in drinking water and on hands has been associated with subsequent diarrhea^41^. Therefore, our findings on the effects of rainfall and temperature on *E. coli* in different types of samples are likely informative of diarrheal risks associated with these pathways under extreme weather conditions.

Bangladesh experiences a high burden of enteric infections; therefore, data on how weather extremes affect infectious disease transmission in this setting can inform other LMICs with similar infection burdens. However, several factors may impact the generalizability of our study.

Bangladesh has a distinct monsoon season that delivers 80% of the annual rainfall; weather effects on environmental contamination may be different in settings with different meteorology and hydrogeology. Also, our study was conducted in a rural environment with high on-premise access to improved water sources and pit latrines but commonly practiced child open defecation and unsafe management of child and animal feces. Weather effects on fecal-oral transmission pathways may vary in urban or densely populated areas and settings with varying WASH infrastructure access and waste management practices.

Our findings provide insight on the causal pathways behind a body of literature that has found increased diarrheal illness during warmer/rainy periods^3,5–7^. As extreme weather events become more frequent and severe, it is important to identify and target the transmission pathways most impacted by these events. Our findings indicate that drinking water and food stored in the home are vulnerable to contamination from extreme weather. Targeting the immediate two-week period, especially the first two days, following extreme rainfall and temperatures to promote interventions and protective practices such as water treatment and safe storage of drinking water and prepared food can reduce exposure to fecal organisms. During the same period following extreme rainfall, avoiding bathing or swimming in surface waterbodies to prevent accidental ingestion and not using surface water for cooking and washing utensils that will contact food or stored water can also help reduce exposures. Our findings provide novel evidence on fecal-oral transmission pathways that are susceptible to weather parameters and time windows for protective measures to interrupt these pathways in the context of climate events. This evidence can inform the effective delivery of interventions to support climate change adaptation and reduce the enteric disease burden associated with weather extremes.

## Data Availability

All data produced in the present study are available upon reasonable request to the authors.

