## Supplementary Materials for "Effects of weather extremes on fecal contamination along pathogen transmission pathways in rural Bangladeshi households"

### Table of Contents

***Table S1***: *E. coli* reporting units and detection limits by sample type **Page 2**

***Table S2:*** Number of samples collected by sample type and study round **Page 2**

***Table S3***: Percentages of study observations within each weather category across different antecedent timeframes **Page 3**

***Table S4***: Number of study observations and geometric mean *E. coli* counts by sample type for each weather category across different antecedent timeframes **Page 4**

***Table S5***: Adjusted *E. coli* count ratios by sample type associated with each weather category across different antecedent timeframes **Page 5**

***Figure S1***: Daily precipitation and temperature, and monthly mean most probable number (MPN) of *E. coli* by sample type across the study period **Page 6**

***Figure S2***: Adjusted *E. coli* count ratios by sample type associated with above-median and elevated temperature, compared to below-median temperature, during different antecedent periods **Page 7**

***Figure S3***: Adjusted *E. coli* count ratios by sample type associated with heatwaves[c] within 7 and 14 days **Page 8**

**Table S1. *E. coli* reporting units and detection limits by sample type**

| | Unit | Lower Limit (MPN) | Upper Limit (MPN) |
|---|---|---|---|
| Stored water | 100 mL | 1 | 2419.6 |
| Food | 1 dry gram | 1–2 [a] | 2,494 – 48,392 [a] |
| Mother hands | 2 hands | 5 | 12,098 |
| Child hands | 2 hands | 5 | 12,098 |
| Source water | 100 mL | 1 | 2,419.6 |
| Soil | 1 dry gram | 1,000–1887 [b] | 2.42 x $10^6$– 4.56 x $10^6$ [b] |
| Ponds | 100 mL | 100 | 241,960 |
| Flies | 1 fly | 100 | 241,960 |

[a] Given a food moisture content range of 3-95%, a lower limit of 1 MPN and upper limit of 2,419·6 MPN per wet gram
[b] Given a soil moisture content range of 0-47%, a lower limit of 1,000 MPN and upper limit of 2,419,600 per wet gram

**Table S2. Number of samples collected by sample type and study round**

| Round | Round 1 | Round 2 | Round 3 | Round 4 | Round 5 | Round 6 | Round 7 | Round 8 | Round 9 | Total |
|---|---|---|---|---|---|---|---|---|---|---|
| Stored water | 1623 | 641 | 592 | 571 | 599 | 581 | 582 | 574 | 587 | 6350 |
| Food | 1646 | 0 | 0 | 329 | 206 | 0 | 0 | 0 | 0 | 2181 |
| Mother hands | 0 | 720 | 705 | 684 | 682 | 668 | 662 | 643 | 633 | 5397 |
| Child hands | 1768 | 720 | 705 | 682 | 673 | 653 | 650 | 626 | 615 | 7092 |
| Source water | 1669 | 0 | 0 | 0 | 0 | 0 | 0 | 0 | 0 | 1669 |
| Soil | 1795 | 0 | 0 | 402 | 341 | 0 | 0 | 0 | 0 | 2538 |
| Ponds | 822 | 0 | 0 | 0 | 0 | 0 | 0 | 0 | 0 | 822 |
| Flies | 610 | 0 | 0 | 0 | 0 | 0 | 0 | 0 | 0 | 610 |
| Total | 9933 | 2081 | 2002 | 2668 | 2501 | 1902 | 1894 | 1843 | 1835 | 26659 |

**Table S3**. **Percentages of study observations within each weather category[a] across different antecedent timeframes. Study observation refers to unique combinations of household GPS coordinates and sample collection dates (total n=7253)**

| | **0 Days** | **1 Day** | **2 Days** | **7 Days** | **14 Days** |
|---|---|---|---|---|---|
| No Rain | 52.4% (3803) | 45.1% (3274) | 41.6% (3014) | 31.3% (2270) | 18.8% (1361) |
| Some Rain | 38.2% (2773) | 39.8% (2888) | 38.3% (2776) | 34.8% (2525) | 34% (2468) |
| Heavy Rain | 4.4% (319) | 7.2% (521) | 8.8% (641) | 11.8% (858) | 14.5% (1051) |
| Extreme Rain | 4.9% (358) | 7.9% (570) | 11.3% (822) | 22.1% (1600) | 32.7% (2373) |
| Below Median Temp | 52.6% (3812) | 48.0% (3483) | 46.1% (3344) | 42.1% (3051) | 38.4% (2785) |
| Above Median Temp | 38.3% (2778) | 40.0% (2903) | 39.9% (2896) | 37.9% (2748) | 34.3% (2486) |
| Extreme Temp | 9.1% (663) | 12.0% (867) | 14.0% (1013) | 20.0% (1454) | 27.3% (1982) |
| No Heatwave | -- | -- | -- | 90.2% (6543) | 87.5% (6343) |
| Heatwave | -- | -- | -- | 9.8% (710) | 12.5% (910) |

[a] Weather classifications are as follows: No Rain (0 mm), Some Rain (<16·4 mm), Heavy Rain (≥16·4 and < 28·2 mm), Extreme Rain (≥ 28·2 mm), Below Median Temp (< 27·1˚C), Above Median Temp (≥27·1 and < 30·2˚C), Extreme Temp (≥ 30·2˚C), No Heatwave (<30·9˚C), Heatwave (≥ 30·19˚C for 3 consecutive days).

**Table S4. Number of study observations and geometric mean *E. coli* counts by sample type for each weather category[a] across different antecedent timeframes**

| | Stored Water (N=6350) | | Food (N=2181) | | Mother Hands (N = 5397) | | Child Hands (N= 7092) | | Source Water (N=1669) | | Soil (N=2538) | | Ponds (N=822) | | Flies (N=610) | |
|---|---|---|---|---|---|---|---|---|---|---|---|---|---|---|---|---|
| | % (n) | Geometric Mean (SD) | % (n) | Geometric Mean (SD) | % (n) | Geometric Mean (SD) | % (n) | Geometric Mean (SD) | % (n) | Geometric Mean (SD) | % (n) | Geometric Mean (SD) | % (n) | Geometric Mean (SD) | % (n) | Geometric Mean (SD) |
| **Day of** | | | | | | | | | | | | | | | | |
| No Rain | 52.8% (3356) | 5.31 (9.95) | 49.4% (1078) | 1.63 (14.94) | 49.2% (2656) | 31.93 (9.88) | 52.3% (3706) | 18.66 (9.24) | 62.1% (1037) | 0.82 (3.90) | 48% (1218) | 121285.80 (13.56) | 75.9% (624) | 4994.44 (5.84) | 56.2% (343) | 432.50 (19.49) |
| Some Rain | 37.8% (2401) | 10.81 (12.10) | 40.4% (881) | 11.81 (27.62) | 39.8% (2146) | 25.17 (9.75) | 38.4% (2722) | 22.41 (10.34) | 33.6% (560) | 1.10 (4.86) | 41.8% (1062) | 139114.00 (10.22) | 19.1% (157) | 4837.13 (6.78) | 39.2% (239) | 1429.36 (23.52) |
| Heavy Rain | 4.3% (270) | 14.77 (12.61) | 5.2% (114) | 41.18 (23.45) | 4.9% (267) | 35.71 (12.90) | 4.4% (313) | 24.14 (9.87) | 2.8% (47) | 1.54 (9.10) | 5.2% (133) | 111340.10 (9.58) | 3.4% (28) | 27879.14 (4.44) | 2.6% (16) | 1544.76 (40.20) |
| Extreme Rain | 5.1% (323) | 21.60 (11.20) | 5% (108) | 30.68 (26.77) | 6.1% (328) | 36.99 (9.26) | 4.9% (351) | 24.72 (8.91) | 1.5% (25) | 1.52 (5.24) | 4.9% (125) | 77025.78 (7.58) | 1.6% (13) | 24471.59 (6.32) | 2% (12) | 369.71 (10.10) |
| Below Median Temp | 52.7% (3345) | 5.70 (10.29) | 62.5% (1363) | 2.09 (17.52) | 43.7% (2360) | 32.88 (9.80) | 52.5% (3721) | 17.68 (9.19) | 79.1% (1320) | 0.90 (4.29) | 61.5% (1562) | 138924.20 (11.58) | 90.5% (744) | 5550.86 (6.09) | 74.4% (454) | 579.08 (21.60) |
| Above Median Temp | 38.4% (2442) | 11.30 (12.25) | 32.2% (703) | 19.48 (28.02) | 44.2% (2384) | 27.23 (9.94) | 38.3% (2719) | 23.39 (10.23) | 20.7% (346) | 1.06 (4.79) | 33% (838) | 97628.63 (10.68) | 9.1% (75) | 4420.02 (7.12) | 25.1% (153) | 1337.50 (22.90) |
| Extreme Temp | 8.9% (563) | 9.96 (10.96) | 5.3% (115) | 32.96 (19.41) | 12.1% (653) | 26.47 (10.51) | 9.2% (652) | 28.07 (9.76) | 0.2% (3) | 0.50 (1.00) | 5.4% (138) | 170818.30 (16.90) | 0.4% (3) | 761.89 (1.66) | 0.5% (3) | 304.56 (22.86) |
| **1 Day** | | | | | | | | | | | | | | | | |
| No Rain | 45.5% (2888) | 4.88 (9.59) | 44.8% (978) | 1.36 (13.63) | 40.9% (2206) | 32.58 (9.72) | 45.1% (3196) | 18.30 (9.33) | 58% (968) | 0.82 (3.90) | 42.9% (1089) | 112160.30 (14.03) | 70.5% (580) | 4893.64 (6.06) | 51% (311) | 408.03 (18.17) |
| Some Rain | 39.6% (2514) | 10.22 (12.26) | 39.5% (861) | 10.21 (26.57) | 41.7% (2252) | 24.99 (9.69) | 39.8% (2824) | 22.32 (10.09) | 33.9% (566) | 1.07 (4.90) | 40.7% (1033) | 159589.20 (10.04) | 21.8% (179) | 5288.68 (5.82) | 42.3% (258) | 1419.65 (25.28) |
| Heavy Rain | 7% (446) | 11.47 (10.91) | 8.2% (179) | 28.51 (23.57) | 8% (432) | 28.96 (11.54) | 7.2% (512) | 21.62 (9.81) | 4.9% (81) | 1.29 (7.07) | 8.2% (207) | 121600.30 (11.03) | 5.4% (44) | 13847.16 (6.23) | 3.8% (23) | 802.28 (30.00) |
| Extreme Rain | 7.9% (502) | 21.15 (11.36) | 7.5% (163) | 38.26 (25.58) | 9.4% (507) | 40.22 (10.46) | 7.9% (560) | 24.83 (9.35) | 3.2% (54) | 1.17 (3.82) | 8.2% (209) | 67865.91 (7.23) | 2.3% (19) | 14685.42 (9.01) | 2.9% (18) | 468.43 (12.00) |
| Below Median Temp | 48.3% (3065) | 5.42 (10.12) | 57.2% (1247) | 1.82 (14.43) | 39.2% (2118) | 32.35 (9.58) | 47.9% (3396) | 17.51 (9.12) | 74.5% (1243) | 0.90 (4.27) | 60% (1421) | 137132.80 (12.01) | 88.2% (725) | 5526.74 (6.10) | 70.7% (431) | 551.34 (20.99) |
| Above Median Temp | 39.9% (2537) | 11.33 (12.13) | 35.7% (778) | 17.73 (30.44) | 45% (2426) | 28.61 (10.22) | 40.1% (2844) | 23.04 (10.32) | 25.2% (420) | 1.04 (4.80) | 36.6% (930) | 107046.10 (10.09) | 11.3% (93) | 4989.69 (6.64) | 28.4% (173) | 1385.66 (23.97) |
| Extreme Temp | 11.8% (748) | 9.71 (11.40) | 7.1% (156) | 26.66 (19.32) | 15.8% (853) | 25.56 (10.15) | 12% (852) | 26.35 (9.49) | 0.3% (6) | 0.50 (1.00) | 7.4% (187) | 134374.80 (16.45) | 0.5% (4) | 458.58 (2.99) | 0.9% (6) | 316.42 (17.45) |
| **2 Days** | | | | | | | | | | | | | | | | |
| No Rain | 41.8% (2655) | 4.79 (9.51) | 43.5% (950) | 1.30 (13.33) | 36.5% (1971) | 33.18 (9.72) | 41.5% (2945) | 17.58 (9.21) | 56.9% (949) | 0.82 (3.90) | 41.5% (1054) | 112622.60 (14.14) | 68.2% (561) | 4715.59 (6.06) | 50% (305) | 389.66 (17.36) |
| Some Rain | 38.3% (2430) | 9.73 (12.24) | 33.7% (735) | 9.83 (26.93) | 41.4% (2234) | 24.82 (9.72) | 38.2% (2712) | 23.95 (10.25) | 28.3% (473) | 1.09 (4.94) | 35.1% (892) | 152067.20 (10.67) | 18.9% (155) | 5134.47 (5.84) | 35.2% (215) | 1325.16 (23.41) |
| Heavy Rain | 8.9% (563) | 11.47 (11.28) | 10.7% (233) | 23.75 (22.31) | 9.3% (502) | 30.18 (11.73) | 8.9% (630) | 18.94 (9.39) | 7.7% (128) | 1.02 (5.28) | 10.6% (268) | 150455.00 (10.22) | 8.5% (70) | 11793.15 (6.45) | 7.2% (44) | 898.60 (34.03) |
| Extreme Rain | 11.1% (702) | 16.70 (11.52) | 12.1% (263) | 23.29 (27.78) | 12.8% (690) | 36.04 (9.89) | 11.4% (805) | 22.98 (9.45) | 7.1% (119) | 1.20 (5.03) | 12.8% (324) | 88017.53 (7.50) | 4.4% (36) | 12008.60 (5.99) | 7.5% (46) | 1704.53 (27.98) |
| Below Median Temp | 46.3% (2940) | 5.32 (10.11) | 54.6% (1192) | 1.71 (14.84) | 37.6% (2027) | 32.31 (9.46) | 45.9% (3258) | 17.26 (9.10) | 71.7% (1197) | 0.85 (4.01) | 53.4% (1355) | 134851.00 (12.33) | 87.1% (716) | 5586.98 (6.10) | 67.5% (412) | 529.80 (20.71) |
| Above Median Temp | 39.9% (2533) | 11.18 (12.02) | 36.2% (789) | 16.67 (30.30) | 44.1% (2382) | 29.02 (10.36) | 40% (2839) | 23.35 (10.33) | 27.4% (457) | 1.18 (5.42) | 37.3% (947) | 112930.10 (9.66) | 12.3% (101) | 4643.53 (6.57) | 30.3% (185) | 1311.68 (23.92) |
| Extreme Temp | 13.8% (877) | 9.93 (11.50) | 9.2% (200) | 23.94 (21.45) | 18.3% (988) | 25.34 (10.00) | 14% (995) | 25.13 (9.47) | 0.9% (15) | 0.64 (2.55) | 9.3% (236) | 122036.30 (16.00) | 0.6% (5) | 803.87 (4.82) | 2.1% (13) | 1406.45 (26.72) |
| **7 Days** | | | | | | | | | | | | | | | | |
| No Rain | 31.3% (1989) | 4.31 (9.35) | 38.8% (847) | 1.23 (13.08) | 24.7% (1332) | 33.58 (9.94) | 31.3% (2217) | 15.17 (8.93) | 51.7% (862) | 0.83 (4.02) | 36.6% (929) | 109019.50 (14.39) | 61.6% (506) | 4740.81 (6.21) | 44% (268) | 386.05 (17.94) |
| Some Rain | 34.9% (2216) | 8.33 (11.11) | 20.3% (442) | 6.73 (24.62) | 41% (2212) | 27.46 (9.76) | 34.8% (2467) | 25.35 (9.63) | 16% (267) | 0.93 (4.58) | 23.2% (589) | 149509.50 (13.03) | 13.6% (112) | 4887.53 (5.28) | 19.3% (118) | 729.95 (19.88) |
| Heavy Rain | 12% (763) | 13.28 (12.47) | 13.8% (300) | 20.18 (22.61) | 12.1% (655) | 29.19 (11.24) | 11.9% (842) | 19.55 (10.06) | 11% (184) | 0.95 (4.37) | 13.7% (348) | 130922.60 (11.03) | 12.3% (101) | 9288.00 (6.31) | 10% (61) | 897.81 (21.09) |
| Extreme Rain | 21.8% (1382) | 12.27 (12.12) | 27.1% (592) | 14.35 (29.24) | 22.2% (1198) | 29.21 (9.67) | 22.1% (1566) | 23.23 (10.25) | 21.3% (356) | 1.20 (5.09) | 26.5% (672) | 126208.70 (7.50) | 12.5% (103) | 6677.73 (6.38) | 26.7% (163) | 1755.19 (28.13) |
| Below Median Temp | 42.4% (2691) | 4.90 (9.77) | 51.4% (1120) | 1.58 (14.39) | 33.6% (1813) | 32.35 (9.31) | 41.9% (2972) | 16.66 (8.94) | 67.5% (1126) | 0.83 (3.93) | 49.9% (1267) | 131726.60 (12.86) | 82.2% (676) | 5538.48 (6.03) | 60.3% (368) | 467.07 (19.83) |
| Above Median Temp | 37.8% (2400) | 10.80 (12.04) | 34.8% (759) | 13.03 (29.78) | 40% (2158) | 30.12 (10.37) | 37.9% (2691) | 23.51 (10.57) | 31.6% (528) | 1.19 (5.36) | 35.8% (908) | 128507.50 (9.14) | 17.2% (141) | 5102.58 (6.80) | 37.5% (229) | 1349.39 (23.96) |
| Extreme Temp | 19.8% (1259) | 11.28 (11.67) | 13.8% (302) | 30.69 (21.65) | 26.4% (1426) | 25.33 (10.15) | 20.1% (1429) | 24.58 (9.33) | 0.9% (15) | 0.64 (2.55) | 14.3% (363) | 97396.32 (13.89) | 0.6% (5) | 803.87 (4.82) | 2.1% (13) | 1406.45 (26.72) |
| No Heatwave | 90.6% (5751) | 7.63 (11.29) | 93% (2028) | 4.43 (25.44) | 86.9% (4689) | 30.99 (10.03) | 90.1% (6392) | 20.42 (9.76) | 100% (1669) | 0.93 (4.40) | 92.5% (2347) | 134597.40 (11.23) | 100% (822) | 5397.41 (6.18) | 100% (610) | 712.12 (22.30) |
| Heatwave | 9.4% (599) | 9.64 (11.64) | 7% (153) | 22.37 (14.93) | 13.1% (708) | 21.11 (9.25) | 9.9% (700) | 21.65 (9.01) | 0% (0) | [b] | 7.5% (191) | 50619.76 (14.25) | 0% (0) | [b] | 0% (0) | [b] |
| **14 Days** | | | | | | | | | | | | | | | | |
| No Rain | 18.8% (1195) | 4.19 (9.63) | 23.4% (511) | 1.05 (12.29) | 14.5% (783) | 35.88 (9.71) | 18.8% (1334) | 14.70 (8.75) | 32.5% (543) | 0.89 (4.59) | 22.7% (576) | 89903.28 (15.62) | 37% (304) | 4239.19 (6.50) | 25.7% (157) | 410.14 (19.49) |
| Some Rain | 34.1% (2164) | 6.35 (10.19) | 19.1% (416) | 2.06 (17.50) | 38.5% (2076) | 28.32 (9.78) | 33.8% (2396) | 23.42 (9.31) | 20.5% (342) | 0.73 (2.97) | 20.7% (525) | 142281.80 (13.69) | 24.3% (200) | 4693.61 (6.00) | 20.7% (126) | 359.45 (16.39) |
| Heavy Rain | 14.7% (934) | 10.00 (11.60) | 19.8% (432) | 9.28 (22.45) | 13.1% (706) | 26.88 (10.55) | 14.5% (1027) | 18.53 (10.72) | 18.5% (308) | 0.89 (3.98) | 19.3% (491) | 117548.00 (12.02) | 19.7% (162) | 8391.78 (5.69) | 17.7% (108) | 672.08 (18.97) |
| Extreme Rain | 32.4% (2057) | 12.40 (12.34) | 37.7% (822) | 14.67 (28.63) | 33.9% (1832) | 29.37 (10.03) | 32.9% (2335) | 22.73 (9.99) | 28.5% (476) | 1.21 (5.41) | 37.3% (946) | 146942.20 (8.18) | 19% (156) | 6536.77 (5.81) | 35.9% (219) | 1612.75 (26.41) |
| Below Median Temp | 38.7% (2458) | 4.67 (9.72) | 49.7% (1083) | 1.51 (14.10) | 29.5% (1590) | 32.37 (9.18) | 38.3% (2714) | 15.99 (8.94) | 65.5% (1093) | 0.82 (3.94) | 47.5% (1205) | 125198.60 (13.20) | 78.8% (648) | 5333.28 (6.03) | 58.7% (358) | 477.95 (19.88) |
| Above Median Temp | 34.3% (2176) | 10.63 (11.81) | 28.6% (624) | 9.38 (27.31) | 36.4% (1966) | 32.89 (10.60) | 34.3% (2433) | 23.40 (10.44) | 27.5% (459) | 1.25 (5.33) | 30.6% (776) | 137487.90 (9.19) | 16.8% (138) | 5539.64 (6.51) | 33.6% (205) | 1043.56 (22.58) |
| Extreme Temp | 27% (1716) | 10.96 (11.74) | 21.7% (474) | 32.72 (24.55) | 34.1% (1841) | 24.17 (9.87) | 27.4% (1945) | 23.47 (9.54) | 7.0% (117) | 0.89 (4.55) | 21.9% (557) | 109283.10 (11.79) | 4.4% (36) | 6057.94 (8.14) | 7.7% (47) | 2803.68 (29.01) |
| No Heatwave | 87.8% (5576) | 7.57 (11.27) | 91.6% (1997) | 4.29 (25.22) | 83.2% (4490) | 31.41 (10.04) | 87.4% (6195) | 20.34 (9.75) | 100% (1669) | 0.93 (4.40) | 90.9% (2308) | 135931.00 (11.21) | 100% (822) | 5397.41 (6.18) | 100% (610) | 712.12 (22.30) |
| Heatwave | 12.2% (774) | 9.64 (11.71) | 8.4% (184) | 24.24 (16.06) | 16.8% (907) | 21.49 (9.37) | 12.6% (897) | 21.91 (9.27) | 0% (0) | [b] | 9.1% (230) | 54121.62 (13.77) | 0% (0) | [b] | 0% (0) | [b] |

a Weather classifications are as follows: No Rain (0 mm), Some Rain (<16·4 mm), Heavy Rain (≥16·4 and < 28·2 mm), Extreme Rain (≥ 28·2 mm), Below Median Temp (< 27·1˚C), Above Median Temp (≥27·1 and < 30·2˚C), Extreme Temp (≥ 30·2˚C), No Heatwave (<30·9˚C), Heatwave (≥ 30·19˚C for 3 consecutive days).

[b] We could not estimate effects of heatwaves on source water (tubewells), ponds and flies due to data sparsity.

### Table S5. Adjusted[a] *E. coli* count ratios by sample type[b] associated with each weather category[c] across different antecedent timeframes

| Adjusted | Stored Water (N=6350) | | Food (N=2181) | | Mother Hands (N = 5397) | | Child Hands (N= 7092) | | Source Water (N=1669) | | Soil (N=2538) | | Ponds (N=822) | | Flies (N=610) | |
|---|---|---|---|---|---|---|---|---|---|---|---|---|---|---|---|---|
| | *E. coli* Ratio (95% CI) | p-value | *E. coli* Ratio (95% CI) | p-value | *E. coli* Ratio (95% CI) | p-value | *E. coli* Ratio (95% CI) | p-value | *E. coli* Ratio (95% CI) | p-value | *E. coli* Ratio (95% CI) | p-value | *E. coli* Ratio (95% CI) | p-value | *E. coli* Ratio (95% CI) | p-value |
| **Day of** | | | | | | | | | | | | | | | | |
| No Rain | ref | -- | ref | -- | ref | -- | ref | -- | ref | -- | ref | -- | ref | -- | ref | -- |
| Some Rain | 1.67 (1.29, 2.17) | < 0.0005 | 1.45 (0.89, 2.35) | 0.357 | 0.96 (0.72, 1.28) | 0.771 | 1.38 (1.09, 1.74) | 0.008 | 0.36 (0.12, 1.07) | 0.066 | 0.87 (0.73, 1.05) | 0.140 | 1.52 (1.03, 2.25) | 0.036 | 0.95 (0.27, 3.39) | 0.941 |
| Heavy Rain | 1.674 (1.02, 2.64) | 0.041 | 2.06 (1.22, 3.49) | 0.007 | 1.62 (1.00, 2.60) | 0.049 | 1.24 (0.68, 2.25) | 0.478 | 1.93 (0.61, 6.13) | 0.266 | 0.74 (0.52, 1.06) | 0.101 | 4.46 (2.42, 8.20) | < 0.0005 | 0.94 (0.30, 2.99) | 0.923 |
| Extreme Rain | 1.98 (1.36, 2.88) | 0.041 | 3.13 (1.63, 5.99) | 0.001 | 0.90 (0.57, 1.41) | 0.632 | 0.92 (0.52, 1.60) | 0.757 | 0.34 (0.07, 1.76) | 0.198 | 0.36 (0.24, 0.53) | < 0.0005 | 3.46 (2.34, 5.11) | < 0.0005 | 0.03 (0.01, 0.19) | < 0.0005 |
| Below Median Temp | ref | -- | ref | -- | ref | -- | ref | -- | ref | -- | ref | -- | ref | -- | ref | -- |
| Above Median Temp | 1.71 (1.39, 2.11) | < 0.0005 | 5.31 (3.23, 8.73) | < 0.0005 | 0.90 (0.70, 1.15) | 0.38 | 1.22 (0.97, 1.54) | 0.095 | [c] | [c] | 0.79 (0.67, 0.93) | 0.004 | [c] | [c] | [c] | [c] |
| Extreme Temp | 1.49 (1.05, 2.12) | 0.025 | 5.60 (3.14, 10.00) | < 0.0005 | 1.02 (0.72, 1.43) | 0.927 | 1.02 (0.65, 1.62) | 0.919 | [c] | [c] | 1.15 (0.82, 1.60) | 0.412 | [c] | [c] | [c] | [c] |
| **1 Day** | | | | | | | | | | | | | | | | |
| No Rain | ref | -- | ref | -- | ref | -- | ref | -- | ref | -- | ref | -- | ref | -- | ref | -- |
| Some Rain | 1.77 (1.37 , 2.29) | < 0.0005 | 1.19 (0.69, 2.07) | 0.534 | 0.97 (0.73, 1.27) | 0.798 | 1.03 (0.80, 1.34) | 0.807 | 0.28 (0.12, 0.68) | 0.005 | 0.95 (0.78, 1.17) | 0.634 | 1.35 (0.93, 1.96) | 0.111 | 0.91 (0.18, 4.75) | 0.913 |
| Heavy Rain | 1.23 (0.81 , 1.87) | 0.324 | 1.49 (0.80, 2.78) | 0.475 | 1.44 (0.94, 2.20) | 0.093 | 0.80 (0.50, 1.28) | 0.345 | 0.58 (0.09, 3.51) | 0.42 | 0.89 (0.68, 1.16) | 0.367 | 2.91 (1.24, 6.82) | 0.014 | 0.64 (0.15, 2.71) | 0.549 |
| Extreme Rain | 2.25 (1.57 , 3.21) | < 0.0005 | 2.13 (1.19, 3.86) | 0.011 | 1.18 (0.84, 1.66) | 0.348 | 0.97 (0.63, 1.49) | 0.877 | 0.10 (0.02, 0.62) | 0.014 | 0.35 (0.25, 0.48) | < 0.0005 | 2.74 (1.51, 4.95) | 0.001 | 0.28 (0.06, 1.39) | 0.119 |
| Below Median Temp | ref | -- | ref | -- | ref | -- | ref | -- | ref | -- | ref | -- | ref | -- | ref | -- |
| Above Median Temp | 1.69 (1.35, 2.10) | < 0.0005 | 11.71 (7.21, 19.02) | < 0.0005 | 0.92 (0.70, 1.21) | 0.554 | 1.25 (0.97, 1.61) | 0.088 | [c] | [c] | 0.89 (0.75, 1.04) | 0.145 | [c] | [c] | [c] | [c] |
| Extreme Temp | 1.62 (1.19, 2.22) | 0.002 | 11.80 (6.48, 21.48) | < 0.0005 | 0.91 (0.66, 1.25) | 0.565 | 0.93 (0.63, 1.37) | 0.723 | [c] | [c] | 0.99 (0.73, 1.34) | 0.959 | [c] | [c] | [c] | [c] |
| **2 Days** | | | | | | | | | | | | | | | | |
| No Rain | ref | -- | ref | -- | ref | -- | ref | -- | ref | -- | ref | -- | ref | -- | ref | -- |
| Some Rain | 1.64 (1.27 , 2.10) | < 0.0005 | 1.06 (0.62, 1.83) | 0.822 | 0.85 (0.65, 1.11) | 0.233 | 1.00 (0.76, 1.32) | 0.991 | 0.20 (0.08, 0.50) | 0.001 | 0.93 (0.74, 1.16) | 0.506 | 1.80 (1.32, 2.45) | < 0.0005 | 5.34 (2.40, 11.89) | < 0.0005 |
| Heavy Rain | 1.21 (0.82 , 1.81) | 0.339 | 1.23 (0.68, 2.22) | 0.49 | 1.24 (0.84, 1.83) | 0.278 | 0.67 (0.43, 1.06) | 0.087 | 0.21 (0.03, 1.62) | 0.135 | 0.86 (0.66, 1.13) | 0.280 | 3.36 (1.71, 6.57) | < 0.0005 | 3.99 (1.64, 9.68) | 0.002 |
| Extreme Rain | 1.88 (1.34 , 2.65) | < 0.0005 | 1.40 (0.81, 2.42) | 0.226 | 0.94 (0.67, 1.32) | 0.717 | 0.88 (0.62, 1.27) | 0.503 | 0.34 (0.01, 1.13) | 0.079 | 0.46 (0.34, 0.61) | < 0.0005 | 2.36 (1.22, 4.57) | 0.011 | 4.10 (1.16, 14.48) | 0.028 |
| Below Median Temp | ref | -- | ref | -- | ref | -- | ref | -- | ref | -- | ref | -- | ref | -- | ref | -- |
| Above Median Temp | 1.58 (1.24, 2.01) | < 0.0005 | 11.35 (6.79, 18.97) | < 0.0005 | 0.99 (0.76, 1.30) | 0.964 | 1.33 (1.01, 1.75) | 0.042 | [c] | [c] | 0.88 (0.74, 1.06) | 0.183 | [c] | [c] | [c] | [c] |
| Extreme Temp | 1.63 (1.20, 2.23) | 0.002 | 13.75 (7.56, 25.00) | < 0.0005 | 0.91 (0.67, 1.24) | 0.555 | 0.98 (0.68, 1.41) | 0.900 | [c] | [c] | 0.94 (0.72, 1.24) | 0.662 | [c] | [c] | [c] | [c] |
| **7 Days** | | | | | | | | | | | | | | | | |
| No Rain | ref | -- | ref | -- | ref | -- | ref | -- | ref | -- | ref | -- | ref | -- | ref | -- |
| Some Rain | 1.42 (1.06 , 1.91) | 0.019 | 0.59 (0.34, 1.02) | 0.059 | 0.72 (0.53, 0.98) | 0.039 | 0.88 (0.61, 1.28) | 0.503 | 0.17 (0.07, 0.45) | < 0.0005 | 1.11 (0.83, 1.49) | 0.494 | 1.14 (0.79, 66) | 0.478 | 1.30 (0.51, 3.32) | 0.582 |
| Heavy Rain | 1.47 (0.96, 2.23) | 0.072 | 0.74 (0.36, 1.54) | 0.426 | 0.96 (0.64, 1.44) | 0.83 | 0.82 (0.53, 1.27) | 0.372 | 0.07 (0.01, 0.47) | 0.006 | 0.84 (0.61, 1.16) | 0.283 | 2.34 (1.22, 4.49) | 0.011 | 0.83 (0.26, 2.60) | 0.744 |
| Extreme Rain | 1.77 (1.22 , 2.57) | 0.002 | 0.81 (0.49, 1.34) | 0.418 | 0.67 (0.48, 0.94) | 0.02 | 1.13 (0.76, 1.68) | 0.548 | 0.13 (0.02, 0.63) | 0.012 | 0.66 (0.49, 0.89) | 0.007 | 1.52 (0.88, 2.63) | 0.135 | 2.62 (0.98, 7.04) | 0.056 |
| Below Median Temp | ref | -- | ref | -- | ref | -- | ref | -- | ref | -- | ref | -- | ref | -- | ref | -- |
| Above Median Temp | 1.61 (1.25, 2.07) | < 0.0005 | 13.09 (8.96, 19.12) | < 0.0005 | 1.10 (0.84, 1.46) | 0.482 | 1.27 (0.92, 1.74) | 0.150 | [c] | [c] | 0.93 (0.77, 1.14) | 0.497 | [c] | [c] | [c] | [c] |
| Extreme Temp | 1.74 (1.33, 2.28) | < 0.0005 | 21.39 (14.46, 31.65) | < 0.0005 | 1.00 (0.76, 1.32) | 0.979 | 0.90 (0.63, 1.27) | 0.533 | [c] | [c] | 0.74 (0.56, 0.97) | 0.029 | [c] | [c] | [c] | [c] |
| No Heatwave | ref | -- | ref | -- | ref | -- | ref | -- | ref | -- | ref | -- | ref | -- | ref | -- |
| Heatwave | 1.15 (0.85, 1.55) | 0.378 | 0.81 (0.47, 1.40) | 0.452 | 0.81 (0.59, 1.11) | 0.189 | 0.79 (0.50, 1.27) | 0.334 | [c] | [c] | 0.54 (0.38, 0.78) | 0.001 | [c] | [c] | [c] | [c] |
| **14 Days** | | | | | | | | | | | | | | | | |
| No Rain | ref | -- | ref | -- | ref | -- | ref | -- | ref | -- | ref | -- | ref | -- | ref | -- |
| Some Rain | 0.85 (0.60, 1.21) | 0.371 | 0.80 (0.53, 1.21) | 0.295 | 0.78 (0.54, 1.15) | 0.210 | 1.05 (0.71, 1.54) | 0.809 | 0.26 (0.09, 0.75) | 0.013 | 1.16 (0.92, 1.45) | 0.204 | 1.15 (0.86, 1.53) | 0.336 | 0.70 (0.36, 1.35) | 0.287 |
| Heavy Rain | 0.81 (0.55 , 1.21) | 0.312 | 0.64 (0.36, 1.14) | 0.132 | 0.87 (0.51, 1.46) | 0.591 | 1.18 (0.73 1.90) | 0.504 | 0.11 (0.02, 0.50) | 0.004 | 0.82 (0.61, 1.10) | 0.182 | 1.99 (1.29, 3.07) | 0.002 | 0.66 (0.28, 1.58) | 0.352 |
| Extreme Rain | 1.04 (0.67 , 1.60) | 0.869 | 0.65 (0.36, 1.15) | 0.141 | 0.81 (0.49, 1.35) | 0.422 | 1.27 (0.82, 1.97) | 0.292 | 0.23 (0.05, 1.07) | 0.062 | 0.78 (0.57, 1.05) | 0.102 | 1.60 (0.99, 2.60) | 0.057 | 1.91 (0.74, 4.90) | 0.179 |
| Below Median Temp | ref | -- | ref | -- | ref | -- | ref | -- | ref | -- | ref | -- | ref | -- | ref | -- |
| Above Median Temp | 1.46 (1.12, 1.91) | 0.005 | 11.37 (7.61, 16.98) | < 0.0005 | 1.23 (0.90, 1.69) | 0.189 | 1.07 (0.76, 1.50) | 0.691 | [c] | [c] | 0.94 (0.74, 1.18) | 0.584 | [c] | [c] | [c] | [c] |
| Extreme Temp | 1.52 (1.17, 1.96) | 0.001 | 20.74 (14.53, 29.61) | < 0.0005 | 1.01 (0.73, 1.39) | 0.944 | 0.78 (0.55, 1.11) | 0.162 | [c] | [c] | 0.80 (0.63, 1.03) | 0.088 | [c] | [c] | [c] | [c] |
| No Heatwave | ref | -- | ref | -- | ref | -- | ref | -- | ref | -- | ref | -- | ref | -- | ref | -- |
| Heatwave | 1.14 (0.86, 1.51) | 0.359 | 1.58 (0.94, 2.66) | 0.086 | 0.79 (0.57, 1.09) | 0.149 | 0.74 (0.51, 1.08) | 0.116 | [c] | [c] | 0.50 (0.36, 0.70) | < 0.0005 | [c] | [c] | [c] | [c] |

[a] Models for rainfall adjusted for temperature and vice versa. All adjusted models also controlled for the following variables: Binary intervention variable (intervention or control), sex of index child, age of index child (in days), number of children under the age of 18 in the household, number of people in the compound, mother's age (in years), mother's educational status, food security category (using HFIAS scale), minutes to water source, household having improved walls, household having improved floors, household wealth quintile based on owned assets, number of cows, goats and chickens/ducks in the compound, source water origin from a tubewell. Food models included hours since stored food was prepared and stored drinking water models included covered storage container, narrow-mouth storage container, and hours water has been stored.

[b] We could not estimate effects of extreme temperature and heat waves on source water (tubewells), ponds and flies due to data sparsity.

[c] Weather classifications are as follows: No Rain (0 mm), Some Rain (<16·4 mm), Heavy Rain (≥16·4 and < 28·2 mm), Extreme Rain (≥ 28·2 mm), Below Median Temp (< 27·1˚C), Above Median Temp (≥27·1 and < 30·2˚C), Extreme Temp (≥ 30·2˚C), No Heatwave (<30·9˚C), Heatwave (≥ 30·19˚C for 3 consecutive days).

**Figure S1: Daily precipitation and temperature, and monthly mean most probable number (MPN) of *E. coli* by sample type across the study period**

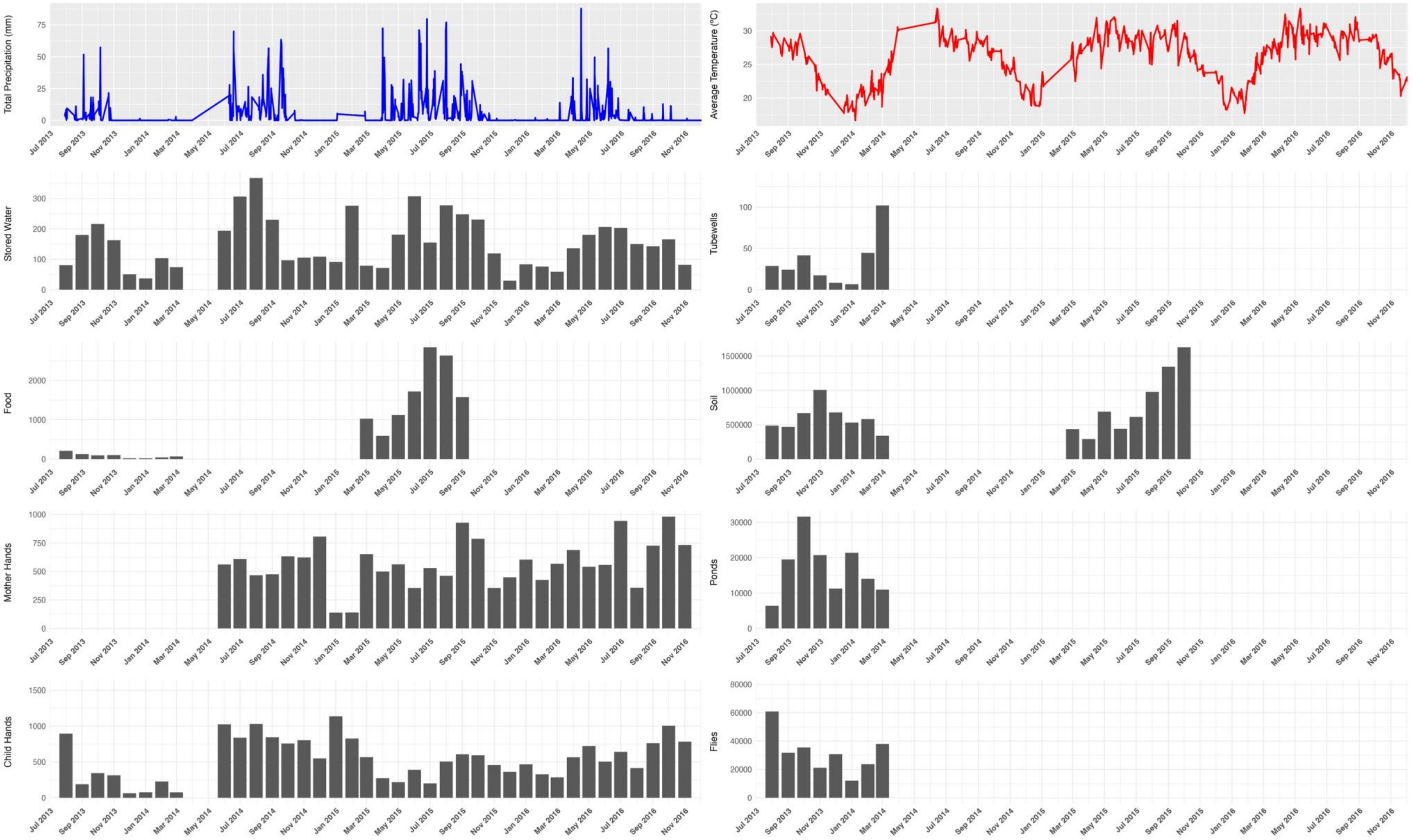

**Figure S2: Adjusted[a] *E. coli* count ratios by sample type associated with above-median and elevated temperature[b], compared to below-median temperature, during different antecedent periods**

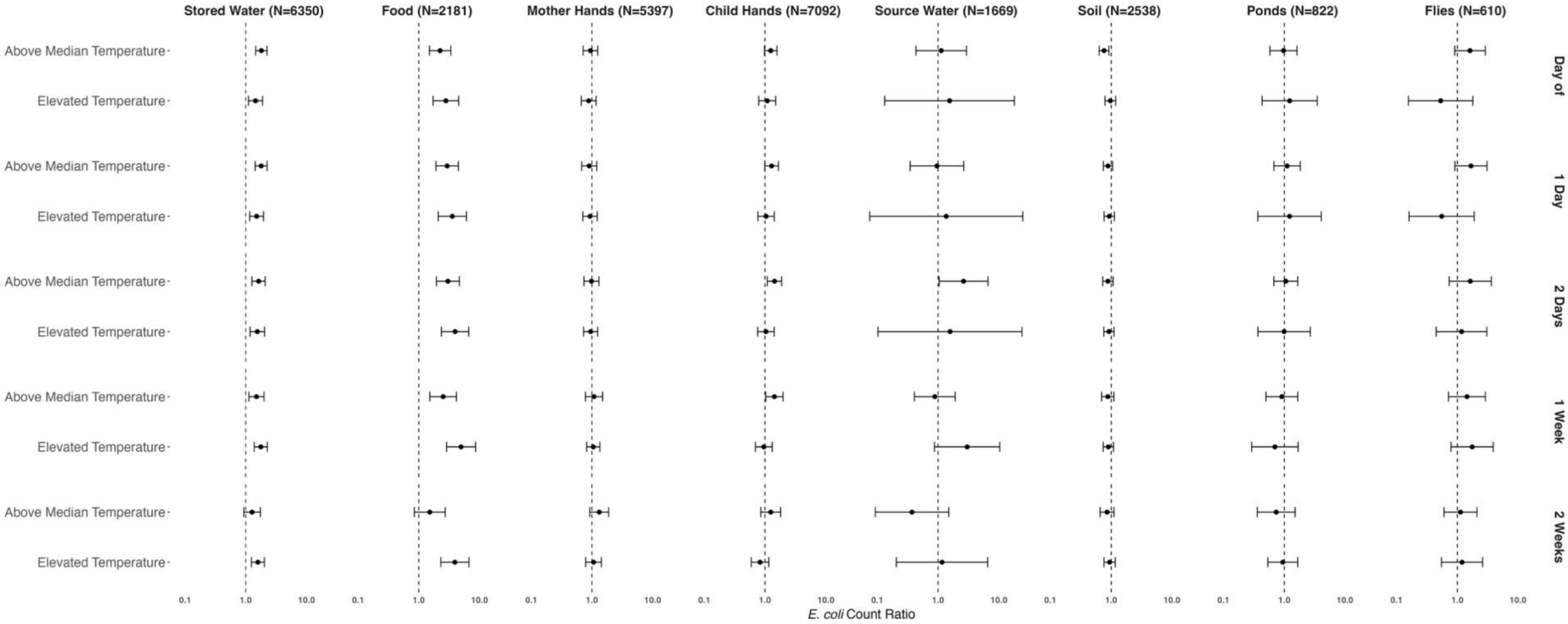


[a] All adjusted temperature models controlled for the following variables: Rolling mean rainfall for the same antecedent period, binary intervention variable (intervention or control), sex of index child, age of index child (in days), number of children under the age of 18 in the household, number of people in the compound, mother's age (in years), mother's educational status, food security category (using HFIAS scale), minutes to water source, household having improved walls, household having improved floors, household wealth quintile based on owned assets, number of cows, goats and chickens/ducks in the compound, source water origin from a tubewell. Food models included hours since stored food was prepared and stored drinking water models included covered storage container, narrow-mouth storage container, and hours water has been stored.

[b] Below Median Temperature (< 27·1˚C), Above Median Temperature (≥27·1 and < 30·2˚C), Elevated Temperature (≥29·3˚C).

**Figure S3. Adjusted[a] *E. coli* count ratios by sample type[b] associated with heatwaves[c] within 7 and 14 days**

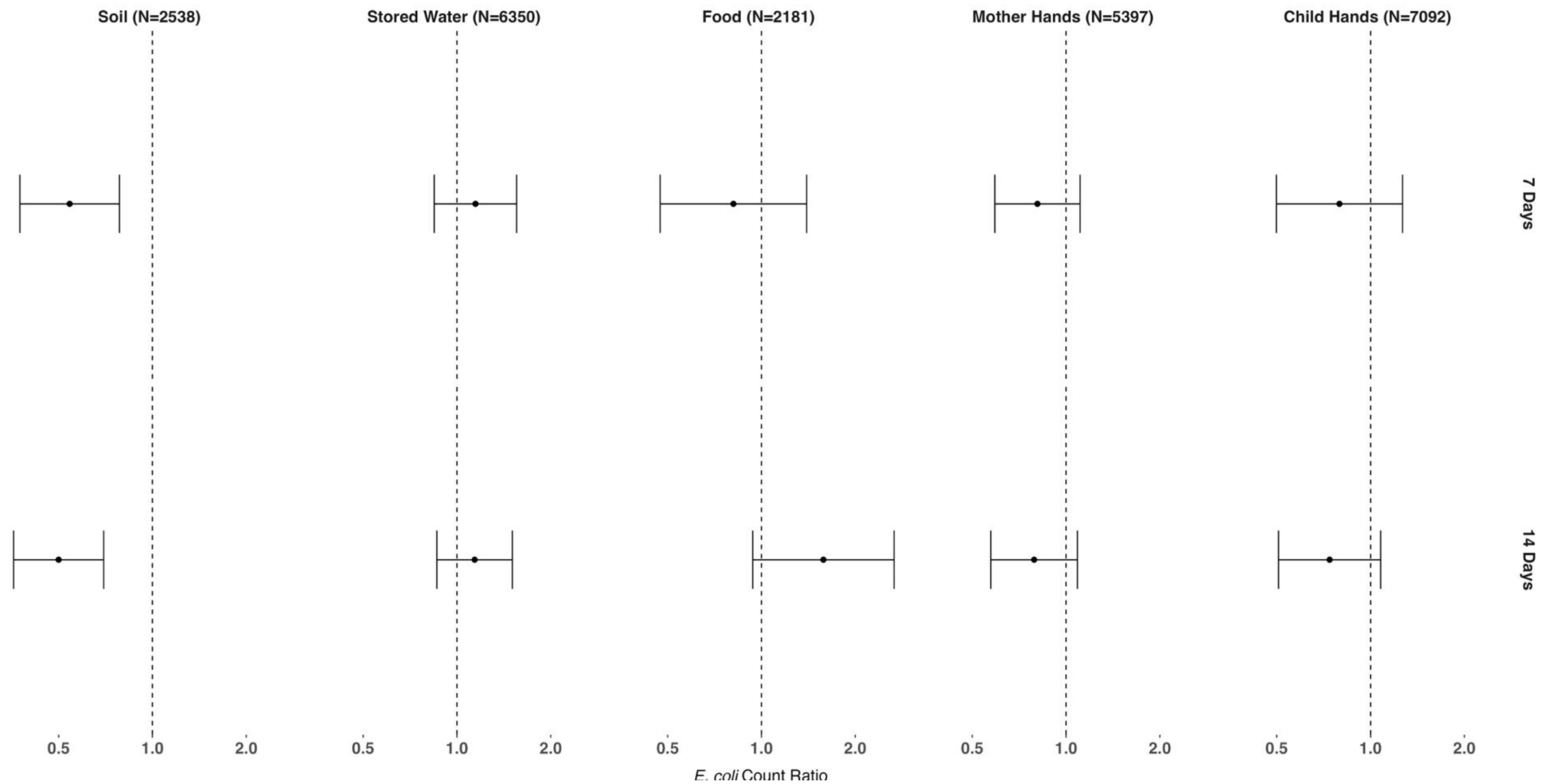


[a] All adjusted temperature models controlled for the following variables: Rolling mean rainfall for the same antecedent period, binary intervention variable (intervention or control), sex of index child, age of index child (in days), number of children under the age of 18 in the household, number of people in the compound, mother's age (in years), mother's educational status, food security category (using HFIAS scale), minutes to water source, household having improved walls, household having improved floors, household wealth quintile based on owned assets, number of cows, goats and chickens/ducks in the compound, source water origin from a tubewell. Food models included hours since stored food was prepared and stored drinking water models included covered storage container, narrow-mouth storage container, and hours water has been stored.

[b] We could not estimate effects of heatwaves on source water (tubewells), ponds and flies due to data sparsity.

[c] Heatwave defined as daily maximum temperature values >95th percentile (30·9°C) for three consecutive days.
